# Polygenic risk, susceptibility genes, and breast cancer over the life course

**DOI:** 10.1101/2020.04.17.20069229

**Authors:** Nina Mars, Elisabeth Widén, Sini Kerminen, Tuomo Meretoja, Matti Pirinen, Priit Palta, FinnGen, Aarno Palotie, Jaakko Kaprio, Heikki Joensuu, Mark Daly, Samuli Ripatti

## Abstract

Polygenic risk scores (PRS) for breast cancer have potential to improve risk prediction, but there is limited information on their clinical applicability. We set out to study how PRS could help in clinical decision making. Among 99,969 women in the FinnGen study with 6,879 breast cancer cases, the PRS was associated not only with breast cancer incidence but also with a range of breast cancer-related endpoints. Women with a breast cancer PRS above the 90^th^ percentile had both higher breast cancer mortality (HR 2.40, 95%CI 1.82-3.17) and higher risk for non-localized disease at diagnosis (HR 2.94, 95%CI 2.63-3.28), compared to those with PRS <80^th^ percentile. The PRS modified the breast cancer risk of two high-impact frameshift risk variants. Women with the c.1592delT variant in *PALB2* (242-fold enrichment in Finland, 263 carriers) and an average PRS (20-80^th^ percentile) had a lifetime risk of breast cancer at 58% (95%CI 50-66%), which increased to 85% (70-100%) with a high PRS (>90^th^ percentile), and decreased to 27% (15-39%) with a low PRS (<20^th^ percentile). Similarly, for c.1100delC in *CHEK2* (3.7-fold enrichment; 1,543 carriers), the respective lifetime risks were 27% (95%CI 25-30%), 59% (52-67%), and 18% (13-22%). Among breast cancer cases, a PRS >90^th^ percentile was associated with risk of contralateral breast cancer with HR 1.66 (95%CI 1.24-2.22). Finally, the PRS significantly refined the risk assessment of women with first-degree relatives diagnosed with breast cancer, i.e. the combination of high PRS (>90^th^ percentile) and a positive family-history was associated with a 2.33-fold elevated risk (95%CI 1.57-3.46) compared to a positive family history alone. These findings demonstrate opportunities for a comprehensive way of assessing genetic risk in the general population, in breast cancer patients, and in unaffected family members.

## INTRODUCTION

In women, breast cancer is the most commonly diagnosed cancer and the leading cause of cancer-related deaths.^1^ Approximately 5–10% of all breast cancers are estimated to develop due to high-impact germline mutations in breast cancer susceptibility genes, with up to 30% due to pathogenic mutations in *BRCA1* and *BRCA2* and with a smaller proportion carrying mutations in other susceptibility genes, such as *PTEN, TP53, CHEK2, PALB2*, and *STK11*.^2^ While pathogenic mutations in *BRCA1* and *BRCA2* are less common in Finns,^3^ two frameshift mutations, c.1592delT (rs180177102) in *PALB2* and c.1100delC (rs555607708) in *CHEK2* have an unusually high allele frequency in Finland, which provides a unique opportunity to explore the impact of these mutations in the population. *PALB2* (Partner and Localizer of *BRCA2*) encodes a key tumour suppressor protein that functions through affecting *BRCA2* nuclear localization and DNA damage response functions, and through interacting with *BRCA1*.^4^ The second gene, *CHEK2* (Checkpoint kinase 2) is a tumour suppressor gene encoding a serine/threonine-protein kinase involved in DNA repair, cell cycle arrest, and apoptosis.^5^

Beyond genetic predisposition caused by high-risk variants in breast cancer susceptibility genes, breast cancer has a highly polygenic mode of inheritance. Large-scale genetic screens have to date identified over a hundred loci associated with risk of breast cancer.^6^ These variants, and many more yet to be discovered, represent common genetic variation acting through a wide range of molecular pathways, in contrast to the rare, high-risk pathogenic variants in high-risk breast cancer susceptibility genes that often disrupt a specific pathway involved in maintaining integrity of DNA repair processes. Individually, the common variants have very small effect sizes, but their cumulative impact in breast cancer risk has been shown to be considerable.^7^ This cumulative effect can be captured in a single measure by a polygenic risk score (PRS), the summed contribution of many common risk variants.^8^ Breast cancer PRS has been shown to improve risk stratification particularly by improving identification of individuals at high risk of breast cancer, and therefore it could potentially serve as a new tool for personalized, risk-based breast cancer screening.^7, 9^

While previous studies have almost exclusively focused on the effect of PRS on overall risk of breast cancer, here we focus on the role of breast cancer PRS across a range of breast cancer-related endpoints and how PRS could help in clinical decision making. We therefore set out to assess three questions with high clinical relevance: 1) how does the PRS impact various breast cancer endpoints such as breast cancer mortality in the population, 2) does the PRS modify the risk of breast cancer in women carrying mutations in the *PALB2* and *CHEK2* genes, and 3) can the PRS inform about outcomes after breast cancer diagnosis and guide risk assessment in relatives of breast cancer patients. We explored these questions using the FinnGen study, which combines nationwide health registries with genomic information and comprises 99,969 women collected across the country, representing 4% of the Finnish adult female population.

## METHODS

### Participants and endpoints

The data comprised of 99,969 Finnish women in the FinnGen, Data Freeze 4. FinnGen comprises prospective epidemiological cohorts (initiated as far back as 1992), disease-based cohorts, and hospital biobank samples (**Supplementary Table 1**). The unique national personal identification number links the genotypes to the Finnish Cancer Registry (available from 1953, with nationwide completeness of solid tumours at 96%^10^), as well as to the national hospital discharge registry (1968-), the registry of surgical procedures (1997-), the national death registry (1969-), and the medication reimbursement registry (1995-). Endpoint definitions were the following: 1) breast cancer cases from the Cancer Registry with diagnosis C50 (International Classification of Diseases for Oncology, 3rd Edition; ICD-O-3) and from the drug reimbursement registry by selecting individuals with a reimbursement code for breast cancer (yielding an additional 856 cases, 847 of whom were diagnosed within 2 years, a delay that sometimes occurs before appearing in the cancer registry) 2) breast cancer mortality through the death registry (C50* (ICD-10) or 174* (ICD-8 and ICD-9) as the underlying or contributing cause of death), 3) non-localized breast cancer at diagnosis from the Cancer Registry, by including individuals with either regional lymph node metastases, distant metastases, or non-localized disease without information on cancer extent, 4) synchronous bilateral breast cancer (from the Cancer Registry), 5) contralateral breast cancer defined as breast cancer in the opposite breast diagnosed over 6 months after the date of the primary breast cancer diagnosis (from the Cancer Registry), 6) mastectomy (procedure codes under ‘HAC’), and 7) breast-conserving surgery (procedure codes under ‘HAB’, conditional on having breast cancer). Out of 6,879 breast cancer cases identified through the endpoint definition 1), detailed Cancer Registry information for endpoint definitions 3) to 5) was available for 6,019 breast cancer cases.

### Genotyping and imputation

FinnGen samples were genotyped with Illumina and Affymetrix arrays (Illumina Inc., San Diego, and Thermo Fisher Scientific, Santa Clara, CA, USA), and genotype calls were made with the GenCall or zCall (for Illumina) and the AxiomGT1 algorithm for Affymetrix data. Individuals with ambiguous gender, high genotype missingness (>5%), excess heterozygosity (+-4SD) and non-Finnish ancestry were excluded; as well as all variants with high missingness (>2%), low HWE P-value (<1e-6) and minor allele count (MAC<3). Array data pre-phasing was carried out with Eagle 2.3.5^11^ with the number of conditioning haplotypes set to 20,000. Genotype imputation was done with Beagle 4.1^12^ (as described in https://dx.doi.org/10.17504/protocols.io.xbgfijw) by using the SISu v3 population-specific reference panel developed from high-quality data for 3,775 high-coverage (25-30x) WGS in Finns.

### PALB2 and CHEK2 variants

Based on a genome-wide association study (GWAS) for breast cancer within FinnGen, we chose two previously reported Finnish-enriched frameshift variants for further analyses, rs180177102 (c.1592delT) in *PALB2* and rs555607708 (c.1100delC) in *CHEK2*. Genotype data batches with an imputation INFO score <0.8 were excluded. This excluded 12,599 women from analyses involving the *PALB2* variant, but no exclusions were needed for *CHEK2. PALB2* mutation carrier status was ignored in analyses involving the *CHEK2* variant, and vice versa. Women homozygous for the *CHEK2* variant were analysed jointly with the heterozygotes.

### Polygenic risk score

We used a previously built and validated breast cancer PRS.^6, 13^ In short, its input weights are from a large independent GWAS,^6^ and LDpred was used to account for linkage disequilibrium among loci.^14^ The PRS showed good calibration for overall risk of breast cancer, breast cancer mortality, and risk of non-localized breast cancer at diagnosis (**Supplementary Figure 1**). To create a PRS independent of the *PALB2* and *CHEK2* variants, we excluded the variants within the *CHEK2* gene ±3Mb, and variants within the *PALB2* gene ±2Mb (**Supplementary Figure 2**). The final variant count for the PRS was 6,023,441. For analysing the effect of PRS in *PALB2* or *CHEK2* mutation carriers, the PRS was divided into the following bins: <20^th^ percentile, 20-80^th^ percentiles (reference category), 80-90^th^ percentiles, and >90^th^ percentile. We chose a threshold at the 90^th^ percentile because it corresponds to a lifetime risk of ≥30%, which guidelines consider as the threshold for high risk.^15^

### Geographic variation

Geographic variation is reported by region of birth (obtained from Statistics Finland) as the proportion of individuals with 1) mutations in the *PALB2* or *CHEK2* variants, and 2) high PRS (>90^th^ percentile). Polygon data for the Finnish map were obtained from GADM (https://gadm.org/data.html).

A population structure-related bias analysis was performed by following the approach described in detail in Kerminen et al.^16^ In brief, the method measures the accumulation of PRS differences between the Western and Eastern subpopulations of Finland using a “random PRS”, made from a randomly chosen set of independent (r^2^ <0.1) variants with minor allele frequency >0.05 that are not associated with breast cancer (breast cancer GWAS^6^ p-value >0.5). If such random PRS accumulated differences between the subpopulations, that could indicate a population genetic bias in effect estimates of the GWAS, rather than a real difference in genetic susceptibility of breast cancer between the subpopulations. We found no evidence of such bias (**Supplementary Figure 3**), which indicates that any detected geographic variation in the PRS is unlikely to result from a population genetic bias.

### Risk assessment in first-degree relatives

The pairs of first-degree relatives were inferred with KING v2.2.4^17^ by a kinship coefficient ranging between 0.177 and 0.354 (inference based on 57K variants). To analyse the impact of family history in first-degree relatives, we randomly chose one female relative for each woman with at least one first-degree relative in the dataset. If both women in the pair were breast cancer cases, we used the year of diagnosis to select the woman diagnosed later as the relative of interest. Each woman could appear only once in the group we inferred the risk for.

### Statistical analysis

The GWAS for breast cancer was run using a generalized mixed model implemented in SAIGE,^18^ adjusting for age, batches, and the first ten principal components of ancestry. We estimated hazard ratios (HRs) and 95% confidence intervals (CI) with the Cox proportional hazards model, and used Schoenfeld residuals and log-log inspection for assessing the proportional hazards assumption. We used age as the time scale, with batches and the first ten principal components as covariates. In the analyses on breast cancer cases only, follow-up started from the diagnosis, and survival over one year was required, as we were interested in assessing long-term survival. Follow-up ended at the first record of the endpoint of interest, death, or at the end of follow-up on December 31, 2018, whichever came first. All tests were two-tailed. Goodness-of-fit for the PRS was assessed with a method proposed by May & Hosmer for a Cox proportional hazards model.^19^ Lifetime risk by age 80 was estimated from the adjusted survival curves, with 95% CIs obtained by normal approximation. The adjusted survival curves were plotted with the R package *survminer*. Statistical interaction was assessed by introducing an interaction term to the survival model, with the PRS on a continuous scale. For statistical analyses, we used R 3.6.1.

Patients and control subjects in FinnGen provided informed consent for biobank research, based on the Finnish Biobank Act. Alternatively, older research cohorts, collected prior the start of FinnGen (in August 2017), were collected based on study-specific consents and later transferred to the Finnish biobanks after approval by Valvira, the National Supervisory Authority for Welfare and Health. Recruitment protocols followed the biobank protocols approved by Valvira. The Ethics Review Board of the Hospital District of Helsinki and Uusimaa approved the FinnGen study protocol Nr HUS/990/2017.

The FinnGen project is approved by the Finnish Institute for Health and Welfare (THL), approval number THL/2031/6.02.00/2017, amendments THL/1101/5.05.00/2017, THL/341/6.02.00/2018, THL/2222/6.02.00/2018, THL/283/6.02.00/2019), Digital and population data service agency VRK43431/2017-3, VRK/6909/2018-3, the Social Insurance Institution (KELA) KELA 58/522/2017, KELA 131/522/2018, KELA 70/522/2019 and Statistics Finland TK-53-1041-17.

The Biobank Access Decisions for FinnGen samples and data utilized in FinnGen Data Freeze 4 include: THL Biobank BB2017_55, BB2017_111, BB2018_19, BB_2018_34, BB_2018_67, BB2018_71, BB2019_7 Finnish Red Cross Blood Service Biobank 7.12.2017, Helsinki Biobank HUS/359/2017, Auria Biobank AB17-5154, Biobank Borealis of Northern Finland_2017_1013, Biobank of Eastern Finland 1186/2018, Finnish Clinical Biobank Tampere MH0004, Central Finland Biobank 1-2017, and Terveystalo Biobank STB 2018001. Analyses of potential geographic bias of PRS were done with THL biobank permission BB2019_44.

## RESULTS

We studied 99,969 women in FinnGen, with the mean age at the end of follow-up 58.5 (inter-quartile range, IQR 45.3-71.8, range 16.0 to 105.7). In FinnGen, 6,879 (6.9%) women have been diagnosed with breast cancer, with mean age at disease onset of 58.7 (IQR 50.6-66.4, range 21.3 to 98.3 years). We first tested for genome-wide association between breast cancer diagnosis and 16,962,023 million genetic variants **(Supplementary Table 2**). There were 17 associated loci, including associations for the well-known frameshift variants rs180177102 in *PALB2* and rs555607708 in *CHEK2*. The allele frequency for rs180177102 (*PALB2*) was 0.0014 (242-fold enrichment compared to non-Finnish non-Estonian Europeans, NFEE^20^), with 263 heterozygote mutation carriers included in the analyses. The allele frequency for rs555607708 (*CHEK2*) was 0.0073 (3.7 times enriched in Finns compared to NFEE), with 1,534 heterozygotes and 9 homozygote individuals in the dataset.

To characterize the impact of the two frameshift variants on the population level, we first explored the geographic distribution in their carrier status. Both the *PALB2* and *CHEK2* mutations had more carriers in Eastern Finland. The allele frequency ranged from close to 0 in Western Finland, to 3.1% for *CHEK2* in South Karelia and 0.8% for *PALB2* in North Karelia (**Figure 1**). This was in contrast with both higher breast cancer incidence and higher PRS observed in Western and Southern Finland.

**Figure 1.**
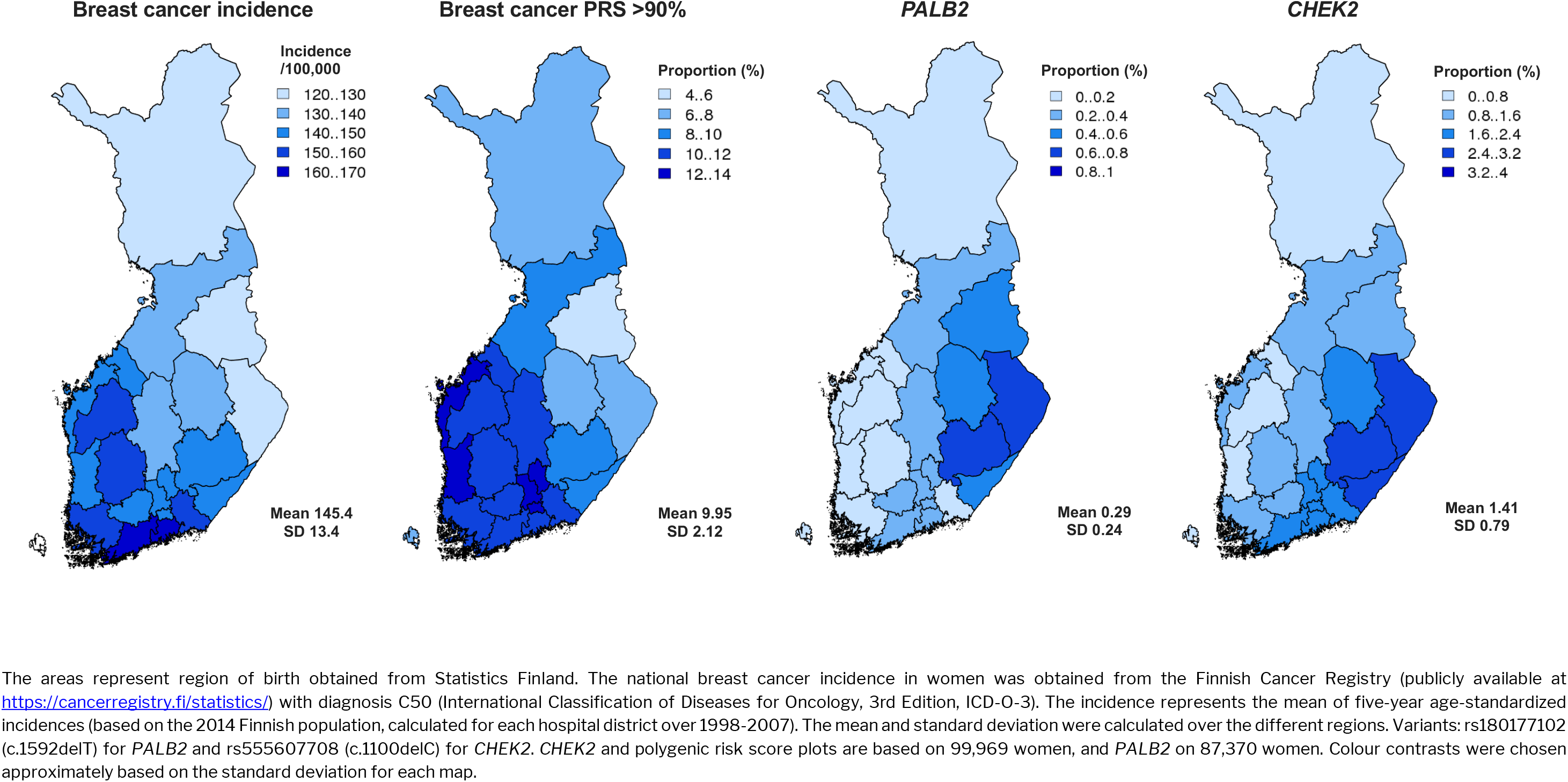
Geographic variation in genetic risk, compared to age-standardized breast cancer incidence. The *PALB2* and *CHEK2* maps show across different regions the proportion of women carrying at least one risk allele for the variants. The proportion of women with the breast cancer polygenic risk score (PRS) above the 90^th^ percentile in each region is estimated with respect to the PRS distribution of the whole country.

### Effect of high-impact variants and polygenic risk in the population

Both variants conferred considerably elevated risk for breast cancer (**Table 1**; detailed counts in **Supplementary Table 3**). The *PALB2* variant conferred a risk increase for breast cancer with a hazard ratio (HR) of 4.80 (95% CI 3.72-6.18, p = 1.08 × 10^−33^), corresponding to a lifetime risk by age 80 of 58.1% (95% CI 52.1-64.1%). The *CHEK2* variant conferred a risk increase for breast cancer with HR 2.12 (95% CI 1.84-2.44), p = 1.33 × 10^−26^), corresponding to a lifetime risk of 31.8% (95% CI 29.5-34.1%). We also identified 92 carriers of five pathogenic *BRCA1/BRCA2* variants (**Supplementary Table 4**) with a 75.1% (95% CI 63.9-86.3%) lifetime risk of breast cancer, but due to the small number of carriers we did not study these variants further. We then compared the risk of elevated PRS to the carriers of *CHEK2* and *PALB2* mutations (**Table 1**). Women with PRS above the 90^th^ percentile had a similar lifetime risk (31.8%, 95% CI 30.9-32.7%) as did *CHEK2* mutation carriers, but a high PRS affected a nearly seven-fold larger group of women.

**Table 1.**
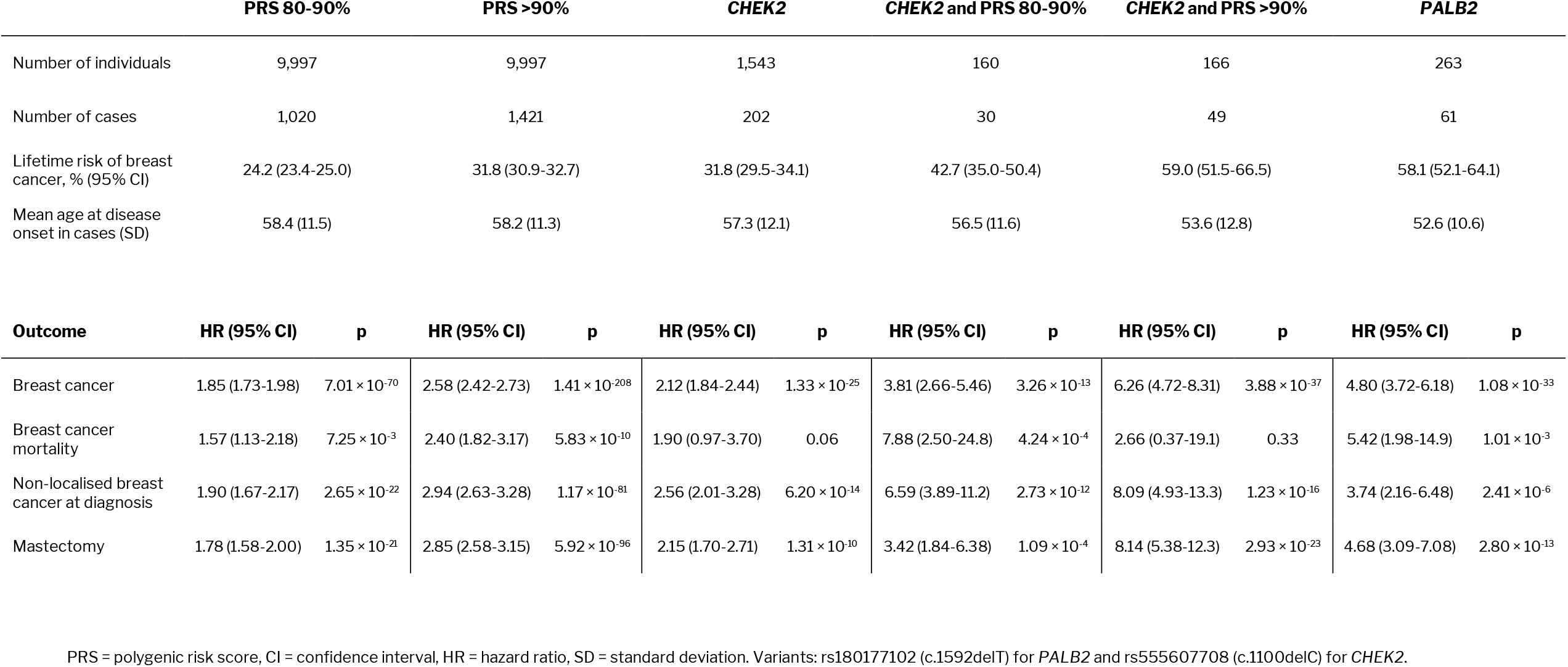
Risk for breast cancer events in the population with different combinations of genetic risk assessment. The reference group for columns with the PRS is individuals with a PRS below the 80^th^ percentile. For columns with *PALB2* or *CHEK2*, the reference group is the rest of the sample.

Of the 6,879 women diagnosed with breast cancer, 31.8% had non-localized breast cancer at diagnosis and 4.6% died of breast cancer by the end of follow-up. Synchronous bilateral breast cancer was diagnosed in 1.6% and contralateral breast cancer in 3.9%. The breast cancer PRS was associated with all of these endpoints in the population (**Table 1**). Compared to individuals with PRS <80^th^ percentile, in women with a PRS between the 80^th^ and 90^th^ percentiles the effect size for an association with non-localized breast cancer at diagnosis was HR 1.90 (95% CI 1.67-2.17, p = 2.65 × 10^−22^), and in the PRS category >90^th^ percentile, HR 2.94 (95% CI 2.63-3.28, p = 1.17 × 10^−81^). Similarly, we observed a strong association with both bilateral breast cancer (PRS 80-90% HR 2.08, 95% CI 1.15-3.76, p = 0.02; PRS >90% HR 4.31 (95% CI 2.73-6.79, p = 3.14 × 10^−10^) and with contralateral breast cancer (PRS 80-90% HR 2.16, 95% CI 1.49-3.15, p = 5.61 × 10^−5^; PRS >90% HR 4.41, 95% CI 3.30-5.89, p = 1.06 × 10^−23^).

Next, we estimated how the PRS modifies breast cancer risk in the mutation carriers. For both *PALB2* and *CHEK2*, a high PRS further increased the breast cancer risk. In terms of absolute lifetime risk for breast cancer by age 80, women with the *PALB2* mutation and average PRS (20-80^th^ percentile) had a lifetime risk of 58.0% (95% CI 50.4-65.6%) which increased to 84.9% (69.9-99.9%) among women with a high PRS (>90^th^ percentile)(**Figure 2**). Accordingly, the lifetime risk decreased to 26.9% (14.7-39.1%) in women with a low PRS (<20^th^ percentile). Women with *CHEK2* and average PRS had a lifetime risk of 27.4% (95% CI 24.5-30.3%), which doubled to 59.2% (51.7-66.7%) in women with a high PRS and decreased to 17.5% (13.0-22.0%) in women with low PRS. We found no evidence of an interaction between the PRS and mutations for neither the *PALB2* variant (p = 0.73) nor the *CHEK2* variant (p = 0.32).

**Figure 2.**
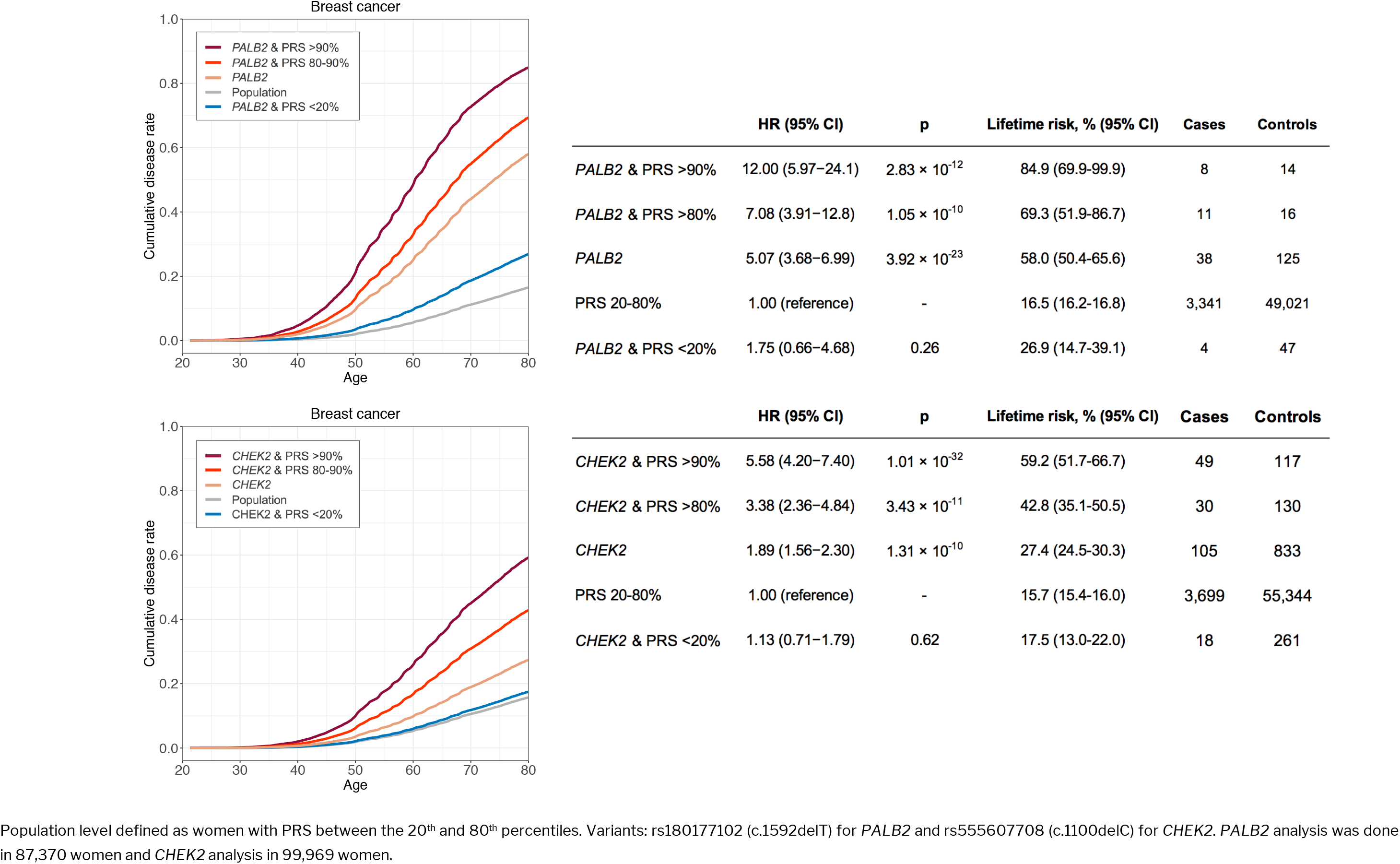
Adjusted survival curves showing how polygenic risk score (PRS) affects the breast cancer risk conferred by the *PALB2* (top) and *CHEK2* (bottom) frameshift mutations.

### Impact of polygenic risk after breast cancer diagnosis

Next, we tested the association between PRS and disease outcomes after the breast cancer diagnosis. A high PRS (>90^th^ percentile) was associated with risk of contralateral breast cancer with HR 1.66 (95% CI 1.24-2.22, p = 0.0007), and the association remained similar when adjusting for clinical factors at baseline (**Table 2**). We found no association between high PRS and breast cancer-related mortality after diagnosis (**Table 2**).

**Table 2.** Impact of high polygenic risk score (PRS) on the risk of contralateral breast cancer and breast cancer mortality in cases.

|  | HR (95% CI) | p |
| --- | --- | --- |
| **Risk of contralateral breast cancer in cases, n = 235** |  |  |
| PRS >90%, unadjusted model | 1.66 (1.24-2.22) | 0.0007 |
| PRS >90%, adjusted model | 1.67 (1.25-2.24) | 0.0006 |
| Age at diagnosis | 1.00 (0.98-1.01) | 0.72 |
| Non-localized breast cancer at diagnosis | 0.89 (0.66-1.19) | 0.42 |
| Breast-conserving surgery at baseline | 1.23 (0.87-1.74) | 0.24 |
| Mastectomy at baseline | 1.21 (0.84-1.75) | 0.30 |
| **Breast cancer mortality in cases, n = 291** |  |  |
| PRS >90%, unadjusted model | 0.93 (0.69-1.24) | 0.61 |
| PRS >90%, adjusted model | 0.89 (0.66-1.20) | 0.44 |
| Age at diagnosis | 1.05 (1.03-1.06) | $2.80 \times 10^{-14}$ |
| Non-localized breast cancer at diagnosis | 2.60 (2.05-3.30) | $4.11 \times 10^{-15}$ |
| Breast-conserving surgery at baseline | 0.52 (0.36-0.74) | 0.0003 |
| Mastectomy at baseline | 1.23 (0.92-1.65) | 0.17 |

HR = hazard ratio, CI = confidence interval. The reference group for the PRS is <80%, and the PRS categories are based on all 99,969 women. Analysis based on 5,979 breast cancer cases, after excluding 40 individuals with follow-up less than one year and cases without detailed Cancer Registry data (mostly newly diagnosed cases; see Methods for details). Controls are breast cancer cases without the event of interest. Mastectomies and breast-conserving surgery at baseline follow the definitions described in the Methods, but were here restricted to procedures $\pm 6$ months from the first breast cancer diagnosis.

### Polygenic risk and breast cancer in first-degree relatives

Lastly, we evaluated how the PRS modifies the risk conferred by a positive family history. This was done by estimating the risk of breast cancer in 9,702 parent-offspring pairs and 9,598 full sibling-pairs. We compared the lifetime risks of A) individuals with a first-degree relative who has both breast cancer and a high PRS (>90^th^ percentile), and B) individuals with a first-degree relative who has breast cancer but does not have a high PRS (<90^th^ percentile) (**Figure 3**). For scenario A), the lifetime risk was 24.6% (95% CI 19.2-30.0%) compared to a lifetime risk of 18.8% (95% CI 16.3-21.3%) in scenario B). If the PRS was known for the woman herself (i.e. the woman we estimate the risk for), the lifetime risk with PRS >90^th^ percentile and a first-degree relative with breast cancer was 34.9% (27.8-42.0%)(**Figure 3**).

**Figure 3.**
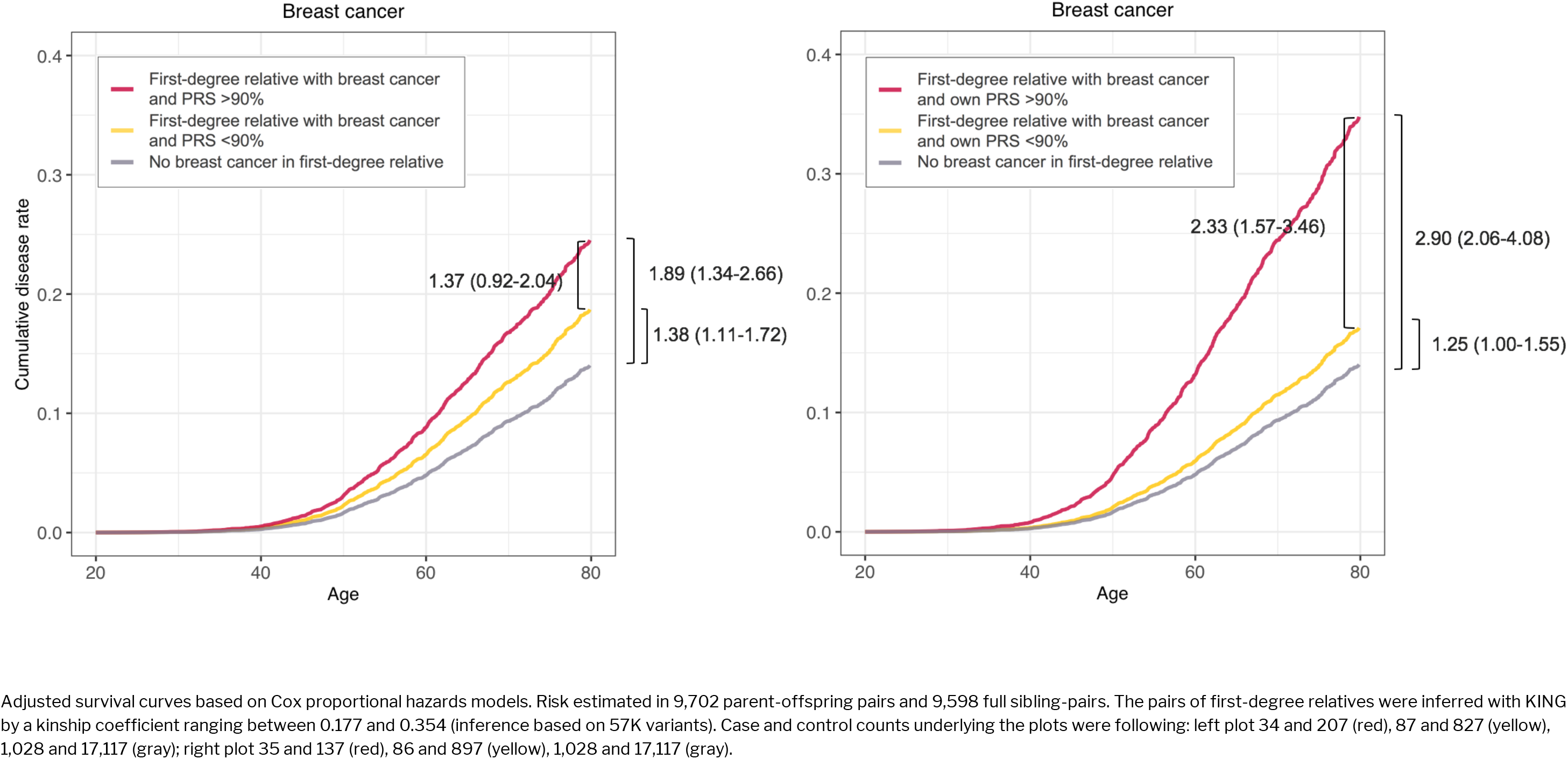
Impact of polygenic risk score (PRS) in estimating the breast cancer risk of women with a first-degree relative diagnosed with breast cancer.

## DISCUSSION

Using large-scale biobank data combining nationwide health registry data with genomic information, we showed that a high breast cancer PRS is associated with breast cancer mortality, more advanced breast cancer at diagnosis, and other breast cancer-related endpoints in the population. We also demonstrate that over the life course, PRS strongly alters the breast cancer incidence in high-impact mutation carriers. After breast cancer diagnosis, individuals with an elevated PRS have an increased likelihood of developing contralateral breast cancer and PRS can considerably improve risk assessment among their female first-degree relatives.

These results allow us to draw several conclusions with clinical implications. First, we show that breast cancer PRS is associated with higher likelihood of non-localized breast cancer at diagnosis and with breast cancer-related mortality in the population. Previous studies have shown that PRS stratifies women for risk of breast cancer^7, 9, 13, 21^ and our results show that this extends also to more advanced breast cancer and breast cancer-related death —this result is important when considering the role of PRS in the context of screening. Harbouring pathogenic mutations in high-risk breast cancer susceptibility genes often prompt intensified medical surveillance and consideration of preventative procedures such as risk-reducing surgery. Our results argue for the need of studies on the impact of targeted actions in women with a high PRS, who currently go undetected. Even after the diagnosis, patients with elevated PRS had a 1.7-fold elevated risk for contralateral cancer, providing additional evidence of increased breast cancer susceptibility, a finding that could warrant intensified or prolonged surveillance in breast cancer cases with elevated PRS. This finding is in line with previous observations that familial factors contribute to the risk of contralateral breast cancer.^22, 23^

Second, PRS strongly alters the risk of breast cancer in *PALB2* and *CHEK2* mutation carriers, substantially increasing the risk of breast cancer in women with a high PRS, and lowering the risk towards the population level in women with a low PRS. Deciding on appropriate surveillance and risk-reduction strategies is a clinical challenge particularly for moderate-risk mutations such as those in *CHEK2*,^27^ and our results show that additional information provided by the PRS could help in these decisions. A combination of breast cancer PRS in the top decile and a mutation in the *CHEK2* variant increased the lifetime risk to 59% – a risk comparable to that seen in *PALB2* mutation carriers – whereas those with a PRS in the bottom quintile had a risk similar to the population level. That PRS modifies the risk in *PALB2* and *CHEK2* mutation carriers supports previous findings suggesting that common genetic variation at least partly explains the widely observed incomplete penetrance of mutations in breast cancer susceptibility genes.^24-26^ This variation is now measurable on an individual level with the breast cancer PRS, which captures a wide range of molecular pathways. This is likely to include genetic determinants of known breast cancer risk factors such as age at menarche or menopause. The interplay of these genetic factors and the PRS remain to be examined in future studies.

Third, the PRS improved risk assessment of first-degree relatives of women with breast cancer. Women with both family history and PRS in the top decile had a 2.3 times higher breast cancer risk compared to women with family history but a PRS below the top decile. Family history is an essential factor guiding screening strategies of family members of breast cancer patients,^15^ and our findings show that PRS could improve the precision of this assessment.

This is the first study to assess the life-course impact of a breast cancer PRS and its joint effect with high-impact risk variants in the two breast cancer susceptibility genes, *PALB2* and *CHEK2*. We studied one clinically relevant pathogenic frameshift variant for each gene, leveraging their considerable enrichment in an isolated population. The lifetime risk estimates for individuals in the top decile of the PRS were comparable to *CHEK2* mutation carriers – both had a lifetime risk of 32%. Comparable lifetime risks were observed also in individuals with both *CHEK2* mutation and high PRS (59% lifetime risk) and those who are *PALB2* mutation carriers (58% lifetime risk). Based on prior studies, we expect the PRS to modify the risks similarly in *BRCA1* and *BRCA2* mutation carriers.^26^ As these genes are relevant in genetic counselling of breast cancer, PRS evaluation would fit in the current lifetime risk-based screening strategies and refine the risk estimation among mutation carriers. The geographical distribution of the PRS also closely followed the distribution of the breast cancer incidence, unlike the geographical distribution of the high-impact variants. The high-impact variants were more common in a late-settlement region, which has passed an internal genetic bottleneck.^28^ How these differences impact any potential regional genomic-based screening strategies warrants further study.

In conclusion, we show that a high breast cancer PRS comes with a comparable risk profile to frameshift mutations in breast cancer susceptibility genes *PALB2* and *CHEK2* and that the PRS strongly modifies breast cancer risk in the mutation carriers. Even after the breast cancer diagnosis, the PRS was associated with breast cancer susceptibility by increasing the risk of contralateral breast cancer, and it considerably improved risk assessment among the patient’s first-degree relatives. These results demonstrate opportunities for a more comprehensive way of assessing genetic risk in the general population, in breast cancer patients, and in unaffected family members of breast cancer patients. Optimization of these strategies in the clinical setting warrant further study.

## Data Availability

The FinnGen data may be accessed through Finnish Biobanks’ FinnBB portal (www.finbb.fi) and THL Biobank data through THL Biobank (https://thl.fi/en/web/thl-biobank).

## Acknowledgements

We would like to thank Sari Kivikko, Huei-Yi Shen and Ulla Tuomainen for management assistance. Following biobanks are acknowledged for collecting the FinnGen project samples: Auria Biobank (https://www.auria.fi/biopankki), THL Biobank (https://thl.fi/fi/web/thl-biopankki), Helsinki Biobank (https://www.terveyskyla.fi/helsinginbiopankki), Biobank Borealis of Northern Finland (https://www.oulu.fi/university/node/38474), Finnish Clinical Biobank Tampere (https://www.tays.fi/en-US/Research_and_development/Finnish_Clinical_Biobank_Tampere), Biobank of Eastern Finland (https://ita-suomenbiopankki.fi), Central Finland Biobank (https://www.ksshp.fi/fi-FI/Potilaalle/Biopankki), Finnish Red Cross Blood Service Biobank (https://www.veripalvelu.fi/verenluovutus/biopankkitoiminta) and Terveystalo Biobank (https://www.terveystalo.com/fi/Yritystietoa/Terveystalo-Biopankki/Biopankki/). All Finnish Biobanks are members of BBMRI.fi infrastructure (www.bbmri.fi). We also thank study participants for their generous participation at THL Biobank and the National FINRISK study. The content is solely the responsibility of the authors and does not necessarily represent the official views of the National Institutes of Health.

The FinnGen project is funded by two grants from Business Finland (HUS 4685/31/2016 and UH 4386/31/2016) and by eleven industry partners (AbbVie Inc, AstraZeneca UK Ltd, Biogen MA Inc, Celgene Corporation, Celgene International II Sàrl, Genentech Inc, Merck Sharp & Dohme Corp, Pfizer Inc., GlaxoSmithKline, Sanofi, Maze Therapeutics Inc., Janssen Biotech Inc).

This work was supported by the Sigrid Jusélius Foundation [to S.R., A.P., M.P., and H.J.]; University of Helsinki HiLIFE Fellow grants 2017-2020 [to S.R.]; Academy of Finland Center of Excellence in Complex Disease Genetics [grant number 312062 to S.R., 312074 to A.P., 312075 to M.D; 312073 to J.K.; 312076 to M.P.]; Academy of Finland [grant number 285380 to S.R, 128650 to A.P., 308248 to J.K., 288509 to M.P., 218068 and 131449 to H.J.]; The Finnish Innovation Fund Tekes [grant number 2273/31/2017 to E.W.]; Foundation and the Horizon 2020 Research and Innovation Programme [grant number 667301 (COSYN) to A.P]; Cancer Foundation Finland sr [to T.M.]; Cancer Society of Finland [to H.J.]; Jane and Aatos Erkko Foundation [to H.J.]. The funders had no role in study design, data collection and analysis, decision to publish, or preparation of the manuscript.

